# Optoacoustic skin mesoscopy opens a window to systemic effects of diabetes

**DOI:** 10.1101/2020.06.29.20142273

**Authors:** Hailong He, Nikolina-Alexia Fasoula, Angelos Karlas, Murad Omar, Juan Aguirre, Jessica Lutz, Michael Kallmayer, Martin Füchtenbusch, Hans-Henning Eckstein, Anette-Gabriele Ziegler, Vasilis Ntziachristos

## Abstract

Diabetes mellitus affects the microvasculature of several organs, including the eyes, heart, brain, kidneys and skin. The skin is the most accessible organ and could offer a window for detecting diabetes-related systemic effects on the microvasculature. However, assessment of skin microvasculature is typically performed based on invasive histological analysis, which is a method that is not well suited for disease monitoring and application to large populations. We introduce ultra-wide band raster-scan optoacoustic mesoscopy (RSOM) for precise, non-invasive assessment of diabetes-related changes in the dermal microvasculature and skin micro-anatomy, resolved with unprecedented sensitivity and detail without the need of contrast agents. Based on this novel contrast, we investigated whether RSOM could impart a new concept in diabetes healthcare, offering characterization of systemic effects of the disease. We applied pretibial RSOM to 72 patients with diabetes, grouped according to disease complications, and 20 age-matched healthy volunteers. We extracted six label-free optoacoustic biomarkers, including dermal microvasculature density, total dermal blood volume and epidermal parameters. We study the effects of diabetes on these parameters as a function of disease severity and find strong statistically significant differences between microvasculature parameters and diabetes progression. We discuss how RSOM label-free biomarkers can lead to a quantitative assessment of the systemic effects of diabetes, complementing the qualitative assessment allowed by current clinical metrics and possibly, in the future, enable a more precise scoring system capturing the gradual evolution of disease.

---

Diabetes mellitus is a complex metabolic disease with increasing worldwide prevalence, leading to several health complications and aggravating healthcare costs ^1,2^. The disease affects the macro- and the microvasculature of several organs, including the heart, brain, lower limbs, retinas, peripheral nerves, kidneys and skin ^1-4^. In the skin, diabetes-induced microvasculature alterations indicate an adverse disease prognosis, as they compromise tissue perfusion and oxygenation, as well as skin, barriers that may lead to cutaneous infections ^3,5-8^, neuropathy with loss of sensation, ulcerations and other comorbidities ^3,5-7^. These microvascular changes may also be indicative of cardiovascular complications such as coronary artery disease (CAD), carotid artery disease and peripheral arterial disease (PAD) ^9-11^ and occur early in the development of diabetes ^3,4,12^. Therefore, assessment of skin microvasculature could lead to a novel way to assess diabetes onset and progression and allow quantification of the true burden of the disease on the vascular system, as opposed to disease course predictions offered by risk factors.

Currently, characterization of diabetes progression in patients is based largely on the qualitative assessment of clinical symptoms based on questionnaires or a scoring system to assess the presence and quality of these symptoms, such as neuropathic pain, light touch perception, sensory testing and others ^5,6^. This assessment is subjective and time-consuming. More importantly, it only evaluates disease progression at infrequent intervals, during which the disease has advanced significantly enough to yield large pathophysiological changes that present as clinical symptoms relating to loss of function and/or pain. Disease manifestations in the skin microvasculature could serve as a means to observe systemic effects of diabetes in a quantitative fashion and possibly lead to finer and more detailed information on the course of the disease based on gradual changes that are not perceivable as clinical symptoms.

Skin is the largest and most easily accessible organ and could serve as a window for diabetes stage. However, routine assessment of epidermal and dermal microvasculature requires a method appropriate for safe, longitudinal, direct and non-invasive measurements. Located under the highly-scattering epidermis, dermal vasculature is not generally accessible to optical microscopy methods, such as confocal or two-photon microscopy ^13,14^. High frequency ultrasound has been reported to assess skin morphology in diabetic patients ^15-18^. However, speckle effects prevent ultrasound from visualizing small vessels of < 100 µm in diameter ^19^ without applying contrast agents (microbubbles), which limits routine application in humans. Hyperspectral imaging (HIS) has also been considered for assessing oxy- and deoxy-haemoglobin concentration in the skin ^20,21^, but this method cannot visualize skin microvasculature because it suffers from low resolution and quantification accuracy caused by variations in skin absorption and scattering properties. Optical coherence tomography (OCT) has evolved to inform on microvasculature by detecting changes in signal intensity due to microcirculation-induced speckle variations ^22,23^. The technique has been employed to assess retinal vasculature in relation to diabetic retinopathy ^24,25^ or measure the epidermal thickness in patients with type I diabetes mellitus ^26^. However, OCT has not been so far shown useful for assessing dermal microvasculature in diabetic skin, possibly due to the highly scattering nature of the epidermis, which limits the imaging sensitivity at dermal layers.

Although OCT could be investigated as a plausible non-invasive method for assessing cutaneous changes in the microvasculature, we consider herein a newer and evidently more powerful method for resolving microvasculature features. Ultra-Wideband Raster Scan Optoacoustic Mesoscopy (UWB-RSOM) has recently shown marked potential to assess vasculature in humans non-invasively and *in-vivo* ^27-30^. In contrast to OCT, which indirectly detects vasculature by small signal variations due to micro-flows, optoacoustic signals are primarily generated by light absorption of haemoglobin within blood vessels, offering high signal-to-noise ratio detection. RSOM has generated never before available highly-detailed cross-sectional images of human skin, which are not accessible with OCT because it is only sensitive to en-face signals. RSOM also offers greater depth penetration than that achieved by OCT, since the image resolution obeys ultrasonic diffraction and is not sensitive to photon scattering. Moreover, the ultrawide-band nature of the signal detected, extending to more than 100 MHz, ensures high-resolution imaging, similar to that achieved by OCT ^27,31^.

We hypothesized that RSOM could detect and quantify skin features that are affected by the progression of diabetes, including dermal micro-vasculature and the epidermal thickness. Consequently, we postulated that we could employ image analysis techniques to detect and quantify RSOM features associated with the stage of diabetes mellitus, as it is currently characterized based on clinical symptoms and comorbidities. In particular, we investigated the relation between microvasculature features and clinical conditions that are indicative of diabetic progression, such as peripheral diabetic neuropathy and macrovascular atherosclerosis, to assess whether significant changes in micro-vasculature are present in association with these comorbidities ^3,8,32^. Successfully accomplishing these goals would introduce a new label-free and portable technology for quantifying systemic disease burden, possibly serving in the future as a portable tool for studying and monitoring disease progression with fine precision, complementing symptom-based assessments.

To test our hypothesis, we employed three-dimensional RSOM skin images of the lower extremities (pretibial area) of 72 diabetic patients and 20 healthy volunteers. The diabetic group comprised patients with previously diagnosed diabetes and no other symptoms, diabetic patients with peripheral neuropathy and diabetic patients with macrovascular atherosclerotic complications. RSOM illuminated the surface of the skin with 532 nm-wavelength and 1 ns-duration pulses and scanned a 120 MHz broadband ultrasound transducer over a 4×2 mm^2^ field of view (FOV, Fig. 1a). All RSOM datasets included in the study passed algorithmically imposed quality control criteria associated with motion and overall scan quality (*see Methods*). Three-dimensional RSOM images were reconstructed over two frequency bands within the 120 MHz bandwidth employed. Band-selected reconstructions implicitly segmented vessels of different sizes; larger vessels (40-150 nm) are seen in the 10-40 MHz band, whereby smaller vessels (<10-40 nm) are seen in the 40-120 MHz band. Vessels seen in the two different bands are color-coded in the rendered images (*red*: larger vessels; *green*: smaller vessels) so that finer vasculature is highlighted in green colour in the presence of larger vessels (see Fig. 1b).

**Fig 1.**
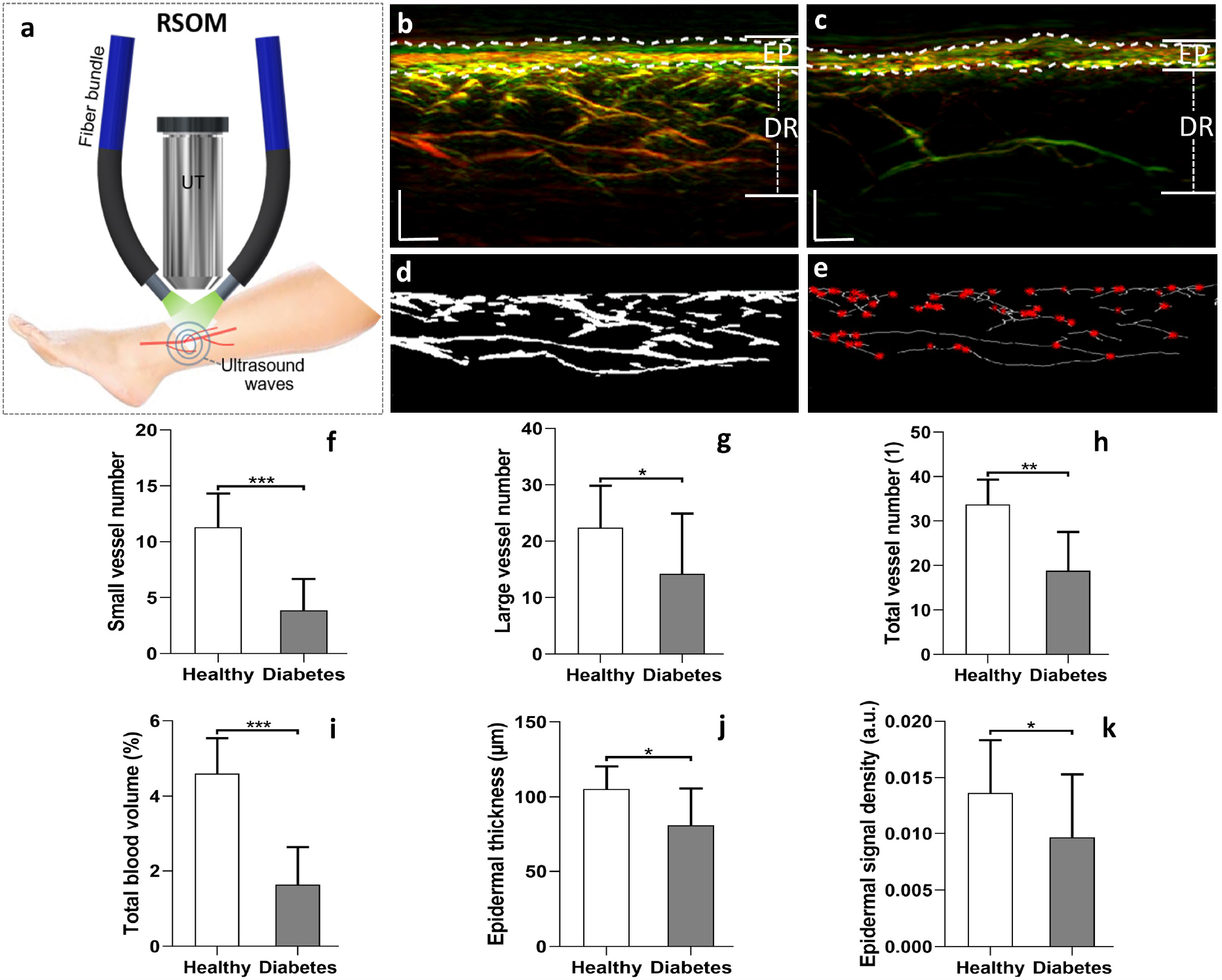
Skin imaging of the lower extremities (distal pretibial region) of healthy volunteers and patients with diabetes using clinical RSOM. **(a)** Schematic of the RSOM system employed for skin measurements, comprising two fibre bundles for illumination and a high frequency ultrasound transducer (UT) that was raster scanned over the skin surface. **(b, c)** Representative tomographic RSOM images of the lower leg of a healthy volunteer **(b)** and a patient with diabetes **(c)**; white dashed lines mark the epidermis (EP) and dermis (DR) layers of the skin. **(d)** Vessel segmentation results from the DR layer of the healthy volunteer. **(e)** Automatic calculation of vessel diameter and numbers of branches; red dots indicate positions of vessel branches, representing vessel numbers. Comparisons between healthy volunteers and diabetic patients: **(f)** the total numbers of small vessels (with diameter <= 40 µm) in DR layer; **(g)** the total numbers of large vessels (with diameter > 40 µm) in DR layer; **(h)** the total numbers of vessels in DR layer; **(i)** the total blood volume of the DR vasculature; **(j)** the average thicknesses of the EP layers, and **(k)** the signal densities of the EP layers. The healthy volunteer group had a population of 20, while the group of diabetic patients had a population of 72. *, **, and *** represent P < 0.05, P < 0.01, and P < 0.001, respectively. Scale bar = 500 µm.

Inspection of 2-band RSOM images of the skin (Fig. 1) visually exemplifies differences between a healthy volunteer and diabetic patient. (Fig. 1b) depicts an image from a 36-year old healthy female volunteer, while (Fig.1c) shows the corresponding image from a 42-year-old male diabetic patient. The images are rendered as maximum intensity projections (MIP) of the entire volume scanned and depict the epidermal (ER) and dermal (DR) layers, reaching a depth of ∼1.5 mm. The typical RSOM appearance of healthy skin shows a dense signal from the epidermis layer and a vascular network in the dermal layer that comprises several blood vessels of various diameters. Conversely, the dermal vessel density is far lower in the diabetic patient compared to the healthy volunteer, a finding that is confirmed by three-dimensional skin visualizations (see Suppl. Fig. S1). Due to the loss of fine dermal vasculature, the diabetic skin exhibits a characteristic high-contrast boundary between the epidermal and dermal layers that is not present in the healthy skin.

To quantify the differences observed by visual inspection of the RSOM images, as well as to extract relevant label-free RSOM biomarkers, we developed two segmentation methods and applied them to the data (see Methods). Briefly, a layer segmentation algorithm based on graph theory and dynamic programming ^33^ identified and separated the epidermis and dermis, as visually marked on the images (c.f. Fig. 1b, Fig. 1c) by the white dash lines. The second method was a vessel segmentation algorithm ^34^, which identified (Fig. 1d) and quantified (Fig. 1e) vascular structures in the dermis layer. Quantification included the computation of the vessel number and the diameter of the different vessels identified. Capitalizing on the broad bandwidth of UWB-RSOM, and based on this image segmentation, we computed and differentially analysed six RSOM image features: (1) the total number of small vessels (NSV, with diameter < 40 µm; 40-120 MHz band) in the dermal layer (Fig. 1f); (2) the total number of large vessels (NLV, with diameter > 40 µm; 10-40 MHz band) in the dermal layer (Fig. 1g); (3) the total vessel number (TVN) in the dermal layer; (4) the total blood volume (TBV) in the dermal layer, computed as ratio of the total number of volume elements (voxels) occupied by the segmented vessels over the total number of voxels in the image; (5) the epidermal thickness (ET), and (6) the epidermal signal density (ESD), computed as the sum of pixel intensity in the epidermis layer and normalized by the total segmented volume of the epidermis layer in the RSOM three-dimensional image.

A next step was to examine the relation of these six metrics to diabetes status (Fig. 1f-i). We found that the mean number of small vessel (Fig. 1f) was about 2.9 times less in diabetic patients than in the healthy volunteers (3.87 ± 2.80 vessels versus 11.3 ± 3.02 vessels). A Mann-Whitney-U-test showed statistically significant differences between the mean number of small vessels in healthy vs. diabetic subjects (P < 0.001). The mean number of large vessels (Fig. 1h) was about 1.6 times less in the diabetic compared to the healthy group (14.21 ± 10.70 vessels versus 22.45 ± 7.42 vessels, P < 0.05). This suggests that the systemic effects of diabetes on vasculature are more prominent in small vessels than in larger vessels. These results were corroborated by analysing the full band RSOM image (Fig. 1h), revealing the total vessel number in the volume examined. The total number of vessels was found to be 18.87 ± 8.70 for diabetic patients vs. 33.75 ± 5.60 for healthy volunteers (P < 0.01).

The total blood volume in the DR layer was also markedly different between the diabetic and healthy groups (Fig.1i), with values of 1.63 % ± 1.23 % and 4.60 % ± 1.21 %, respectively. The Mann-Whitney-U-test here also showed significant differences between the healthy and diabetic subjects (P < 0.001). Analysis of the EP layer also demonstrated statistically significant changes between the two groups. The mean values of EP thickness were 105.29 ± 15.12 µm for healthy volunteers and 81.03 ± 24.61 µm for diabetic patients (Fig.1j) with P < 0.05. Likewise, the signal density of the EP layer (Fig.1k) in the reconstructed segmented volume, which contains contributions from melanin and capillaries, was markedly lower in the diabetic patients than in the healthy volunteers (P < 0.05).

A further unknown of primary interest in this interrogation was the relation of diabetes severity to microvasculature and to the extracted RSOM label-free biomarkers. Diabetes is a chronic disease with systemic effects that generally evolve with time. Therefore, the question of understanding severity is perhaps more critical than the separation of diabetic from healthy groups. Indeed, whereas a diagnostic test separates a continuum of disease stages to healthy and patients based on a threshold value, we see the RSOM role being more closely related to quantifying benchmarks or features associated with different diabetic stages. To address this question, we first examined RSOM features obtained from scans of patients with diabetes without complications and scans from diabetes patients with neuropathy of different severities. The severity of diabetic neuropathy was clinically evaluated using the Neuropathy Disability Score (NDS) and the Neuropathy Symptom Score (NSS) ^35,36^. We divided the patient group into three categories: those with no complications (NC, n = 22), those with low score neuropathy (LN, n = 13; 1≤ NDS ≤ 5 or 1≤NSS ≤ 5) and those with high score neuropathy (HN, n = 12; NDS > 5 or NSS > 5). Representative RSOM images from the healthy group and the three patient groups are depicted in cross-sectional (sagittal) views (Fig. 2a-d) and coronal views (images parallel to the RSOM scan plane) from the epidermal (Fig. 2e-h) and dermal (Fig. 2i-l) layers. The images confirm a reduced vascular density in the dermal layer with progression of the disease. Moreover, observation of the coronal views of the EP layer depict clearly resolved superficial skin ridges in the healthy skin, which change into an amorphous pattern without ridge definition depending on disease status, especially for the neuropathy groups.

**Fig 2.**
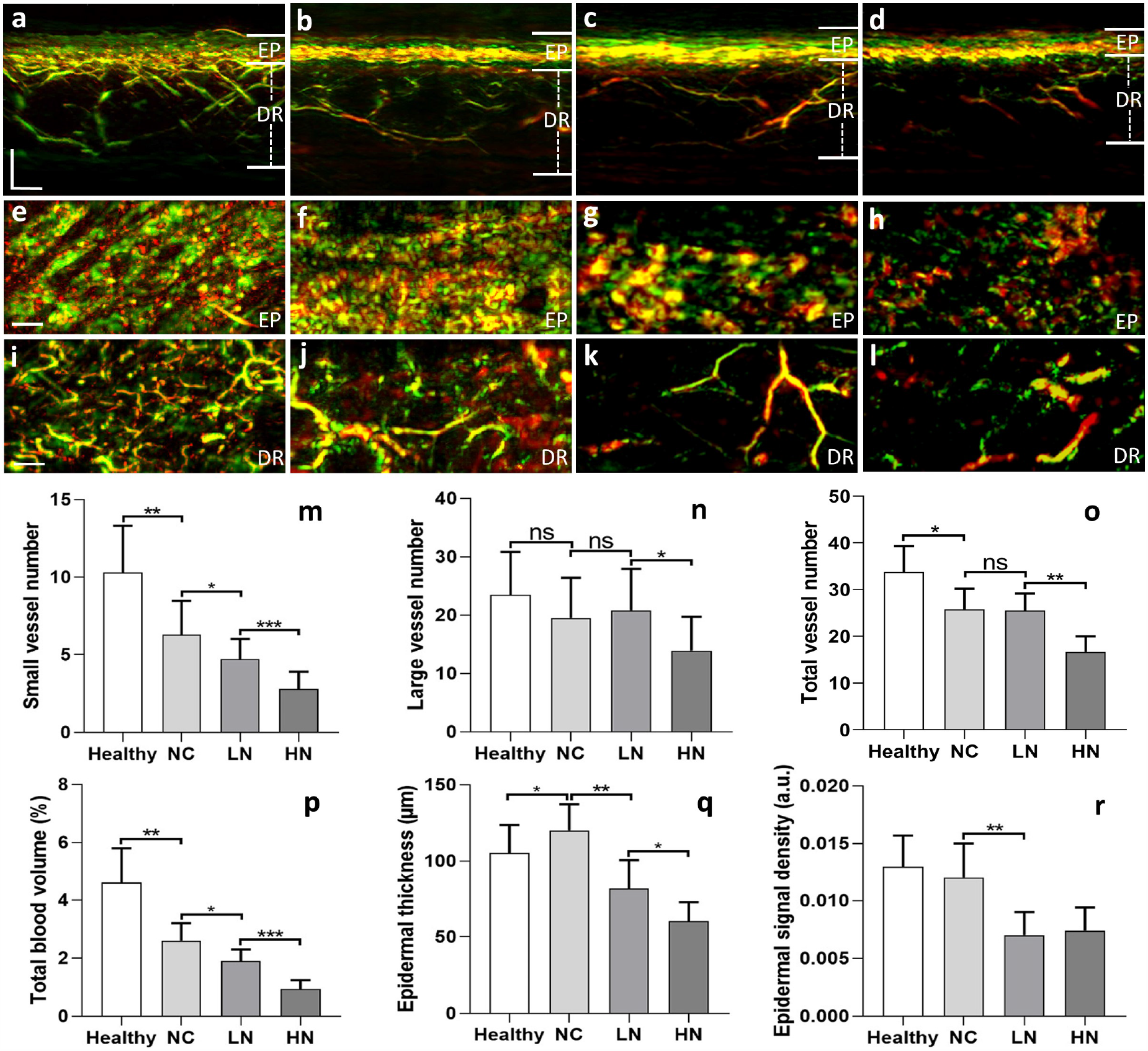
Quantification of RSOM features related to diabetic neuropathy. Patients were grouped as follows: NC (n = 22), diabetic patients with no complications; LN (n = 13), diabetic patients with low score neuropathy (1≤ NDS ≤ 5 or 1≤NSS ≤ 5); HN (n=12), diabetic patients with high score neuropathy (NDS > 5 or NSS > 5). **(a)** Sectional RSOM image of healthy skin. **(b)** Sectional RSOM image of skin from a diabetic patient without neuropathy. **(c)** Sectional RSOM image of skin from a diabetic patient with low score neuropathy (NDS: 3, NSS: 3). **(d)** Sectional RSOM image of skin from a diabetic patient with high score neuropathy (NDS: 9, NSS: 9). MIP images of the EP and DR layers in the coronal views corresponding to (a-d) are displayed in (e-h) and (i-l) respectively. Comparisons amongst the six groups: **(m)** the total number of small vessels (with diameter <= 40 µm) in DR layer; **(n)** the total number of large vessels (diameter > 40 µm) in DR layer; **(o)** the total vessel numbers in DR layer; **(p)** the total blood volume in DR layer; **(q)** the thickness of EP layer and **(r)** the signal density of EP layer as a function of group studied. *, **, and *** represent P < 0.05, P < 0.01, and P < 0.001, respectively. Scale bar = 500 µm. ns=not statistically significant

Quantitative comparisons of the performance of different RSOM features are presented in (Fig. 2f-i). Overall, the count of small vessels (Fig. 2m) demonstrated statistically significant differences between the three patient groups, i.e. healthy versus NC (P < 0.01), NC versus LN (P < 0.05), and LN versus HN (P < 0.001). As shown in (Fig. 2m), the neuropathy grade is visible in the count of small vessels (< 40 µm diameter). However, the presence of neuropathy had no significant effects on the number of large vessels (Fig. 2n). This finding is reflected also in the total vessel number between the healthy and the NC (P < 0.05), as well as between the LN and HN groups (P < 0.01). No marked changes were observed between the NC and LN groups. The computation of blood volume (Fig.2p) demonstrated a similar performance to the small vessel count, with the NC group exhibiting significantly lower blood volume compared to healthy group (P < 0.01). The TBV was further reduced in the LN compared to NC group (P < 0.05). A significant difference in TBV was observed between the LN and HN groups (P < 0.001).

We were also interested in studying the physical manifestations of atherosclerosis in RSOM features. Most of the diabetic patients with atherosclerosis enrolled in this study had also been diagnosed with neuropathy. Therefore, diabetic patients were divided into two groups: diabetic patients with neuropathy and no atherosclerosis (N_NA, n = 25), and diabetic patients with neuropathy and atherosclerosis (N_A, n = 24). Representative cross-sectional (sagittal) views and coronal views of the DR layer from the two groups (Fig. 3a-d) showed marked differences. We saw significant differences in the comparisons of the small, large and total number of vessels between the group with and the group without atherosclerosis (Fig. 3e-g). The small vessel count again exhibited the most statistically significant difference (P < 0.001) between the two groups, compared with the total vessel count (P < 0.01) and large vessel count (P < 0.05). In addition, the total blood volume (Fig. 3h) was significantly reduced in atherosclerotic patients (P < 0.001). Conversely, atherosclerosis had no apparent effect on the EP thickness (Fig. 3i) or the optoacoustic signal density of the EP layer (Fig. 3j).

**Fig 3.**
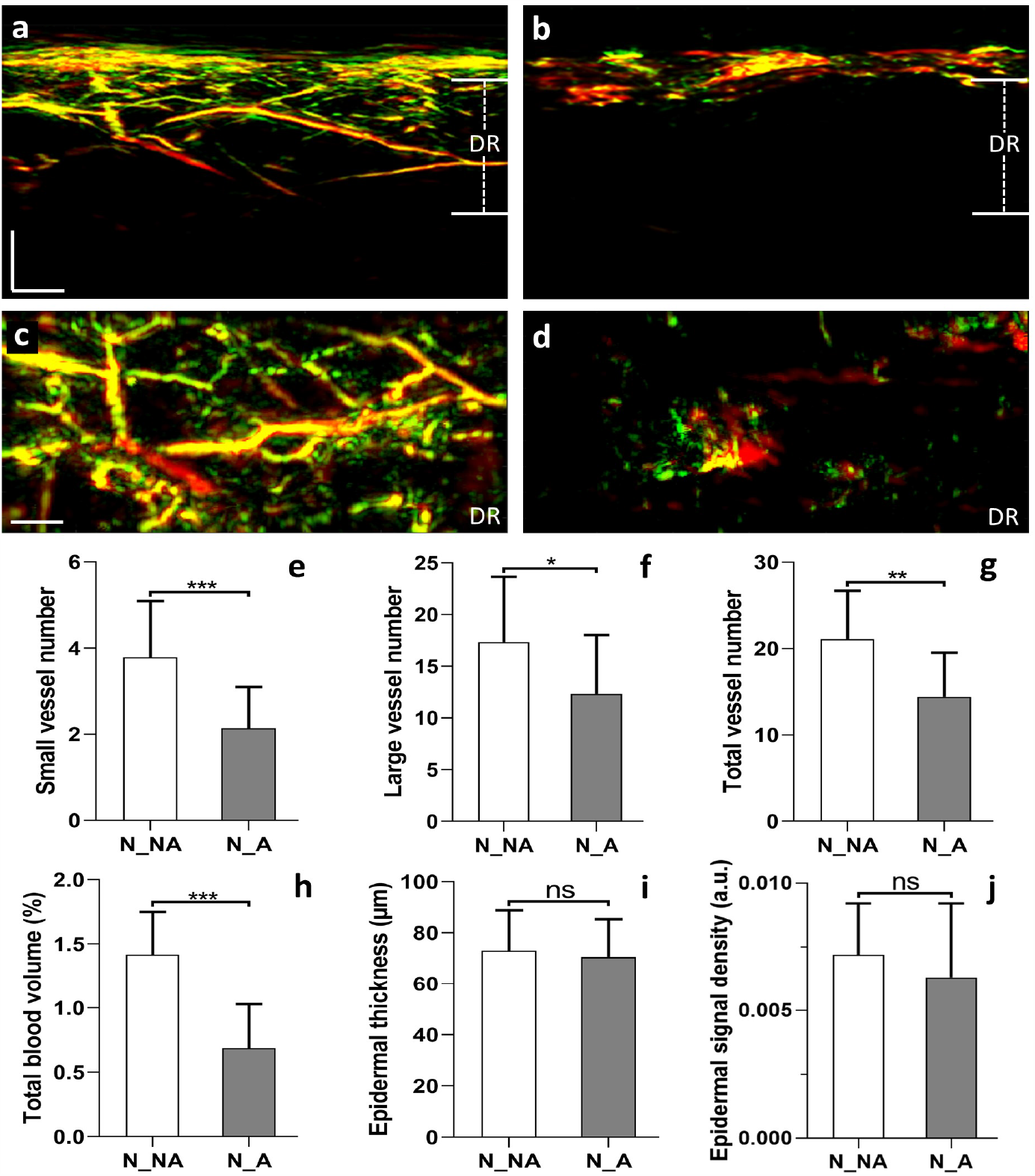
Quantification of RSOM features relating to diabetic neuropathy and atherosclerosis. Patients were grouped as follows: N_NA (n = 25), diabetic patients with neuropathy but no atherosclerosis; N_A (n=24), diabetic patients with neuropathy and the presence of atherosclerosis. **(a)** Sectional RSOM image from N_NA patient. **(b)** Sectional RSOM image from N_A patient. (**c, d**) MIP images of vascular maps in the coronal views of DR layers corresponding to **(a-b)**. Comparisons between the two groups: **(e)** the total number of small vessels (with diameter <= 40 µm) in DR layer; **(f)** the total number of large vessels (with diameter > 40 µm) in DR layer; **(g)** the total vessel numbers in DR layer; **(h)** the total blood volume in DR layer; (i) the thickness of EP layer and **(j)** the signal density of EP layer. *, **, and *** represent P < 0.05, P < 0.01, and P < 0.001, respectively. Scale bar = 500 µm. ns=not statistically significant.

## Discussion

The skin has been heralded as a window for assessing systemic health conditions. This premise has so far held true for identifying a number of conditions, for example infant jaundice or manifestations of systemic sclerosis or fibrosis, based on superficial features appearing on the upper layers of the epidermis. In this work, we employ RSOM as a novel technique that can provide high-resolution imaging under the skin surface and a detailed assessment of dermal micro-vasculature and other skin features. RSOM is the only technique available that can non-invasively provide highly detailed cross-sectional images of optical contrast through the entire skin depth. In this sense, in contrast to the traditional superficial inspection of the human skin, we propose to employ the three-dimensional appearance of the skin to assess systemic effects of diabetes.

Based on histology studies, it is known that diabetes alters human skin microvasculature, reflecting a systemic effect of the disease. These alterations manifest early in the disease development ^5,7,8^. However, the relation of skin microvasculature and diabetes has not been previously investigated in a non-invasive manner, due to the absence of tools that are capable of detailed assessments of fine vasculature. Herein, the ultra-wide band nature of RSOM enables the study of vessels with diameters ranging from less than 10 μm to more than 150 μm, allowing a detailed understanding on the relation between disease severity and vessel size, which was not previously possible. The implications of this RSOM capacity in diabetes are multi-faceted, since successful application of the technology could improve the study of the systemic effects of diabetes and monitor lifestyle or other interventions in a detailed, quantitative way that is not available today. RSOM can play a very different role than blood glucose measurements. While the latter assess the day-to-day glucose status and are necessary for reducing hypoglycaemic incidents, RSOM could monitor an actual state of diabetes using a measure of systemic damage. Importantly, cutaneous microvasculature changes could be non-invasively monitored by RSOM in intervals that are more frequent than the appearance of new clinical symptoms, thus providing a more precise measure of diabetes progression.

We have previously demonstrated that RSOM can provide detailed images of skin vasculature, and that quantitative information can be extracted from these images in relation to dermatology conditions ^27,37,38^. However, it was unknown whether RSOM had the sensitivity to capture diabetes-related changes within skin microarchitecture. Furthermore, the relation between diabetes severity and microvasculature was also unknown. The study herein provided RSOM images of the skin of patients with different diabetes conditions (diabetes severity). Six RSOM label-free biomarkers were extracted from these images: three associated with dermal micro-vasculature (total vessel count, vessel count for vessels < 40 μm in diameter, and vessel count for vessels > 40 μm in diameter), as well as bulk measurements, such as the total blood volume and the thickness and signal density of the epidermis. Visual inspection of RSOM images revealed changes in the patterns observed in the different pathologies examined. Qualitatively, it was generally visible that as diabetes progresses, the vascular density in the dermal layer decreases while the epidermis becomes thinner and less light absorbing. P-tests performed on features quantitatively extracted from the RSOM images statistically confirmed that all these biomarkers are associated with aspects of diabetic progression. Moreover, these analyses identified the small vessel count to be the most indicative marker of diabetes conditions, providing the starkest contrast between the different groups of patients. In other words, the study identified the small vasculature as the component of skin that is most affected by diabetes progression, highlighting the vulnerability of small vessels to systemic effects caused by diabetes and possibly suggesting a diabetes severity label-free biomarker.

A possible route based on the findings herein is that RSOM features could be used to complement diabetes staging. Neuropathy is the most prevalent microvascular complication arising from the disease, and previous studies on skin biopsies have reported changes in the morphology of microvessels correlating with the clinical severity of the case ^39,40^. We have shown here with RSOM a reduction in vessel number, blood volume and epidermal thickness going from patients with no, to mild, to severe neuropathy. In addition to neuropathy, we found a correlation between the presence of macrovascular atherosclerotic disease and a decrease in dermal vessel number and blood volume. These results point to trends that can capture gradual changes of microvasculature alterations indicative of disease progression.

RSOM represents a paradigm shift in the non-invasive evaluation of skin vasculature, well beyond the current state-of-the-art. Depending on the wavelength employed, the method can penetrate several millimetres under the skin surface. Using 532 nm in this study, we focused on visualizing the first millimetre of the skin while retaining high contrast due to the relatively high-absorption of haemoglobin in the green region. Highly detailed RSOM images were showcased herein both as cross-sectional images and as coronal images from different layers. No other method today can achieve this kind of imaging detail and depth, using label-free operation. Moreover, an RSOM system is cost efficient and can be made highly portable to allow disseminated use. Therefore, the results point to the use of RSOM as a highly potent strategy for offering a quantitative assessment of the systemic effects of diabetes and possibly in diabetes staging. Other tests that can measure skin changes include simple visual assessment of the skin surface or Doppler imaging ^41^ to assess changes in blood flow in the skin due to stimuli, such as the post-occlusive increase of shear stress, hyperthermia, or drug applications ^42,43^. Optical Coherence Tomography also offers partial views of skin microvasculature ^44,45^. However, none of these methods offers the quality and detail of RSOM, and consequently, none of these methods have been considered for assessing skin microvasculature and diabetes-related alterations.

There are several reports that microvascular changes occur early in diabetes progression ^3,8^. This observation points to a future prospective study, in which RSOM could be employed in high-risk populations to examine the relation of RSOM biomarkers to the progression from pre-diabetes to diabetes. Such a study could further expand the possible applications of RSOM, not only as a tool to stage and monitor the progression of diabetes, but also as a means of early detection. Although RSOM successfully distinguished between diabetic and healthy populations, the crude separation of a progressing disease with a simple threshold has been noted as a sub-optimal strategy for prevention and diabetes healthcare. This is because people just below a diagnostic threshold are deemed “healthy” and often do not receive the care of patients just above this threshold. It has been argued that it would be better that patients are not separated by a threshold but rather assigned a score so that individuals are better alerted to their condition, monitored closely and be considered for a prevention program ^46,47^. A non-invasive portable and label-free technology such as RSOM could play a vital role in offering quantitative monitoring in high-risk populations and evaluate possible interventions. Although such strategies apply primarily to diabetes 2 patients, the overall need to improve diabetes staging has been outlined for both diabetes 1 and 2 ^46,47^.

In summary, we presented the first optoacoustic mesoscopy images of diabetic patients and non-invasively studied the systemic effects of diabetes on skin microvasculature and skin microanatomy in patients with diabetes mellitus. RSOM allowed six label free biomarkers associated with skin morphology and microvasculature and identified fine vasculature to be the feature most sensitive to diabetes progression. This finding further supports the hypothesis that RSOM could be used as a point-of-care device for quantifying systemic effects of diabetes and providing a quantitative score indicative of diabetic stage. Due to its safety, portability and low cost, quantitative and informative nature, high image quality and ability to quantify label-free biomarkers, RSOM may offer a paradigm shift in the clinical characterization of diabetes, assessing interventions and in prevention programs.

## Methods

### RSOM imaging system

The present study used an in-house portable RSOM imaging system featuring a transducer with a 10-120MHz bandwidth and central frequency at ∼ 50 MHz (Fig. 1a), which has been described in detail elsewhere ^27,48^. Illumination was provided by a pulsed laser at a wavelength of 532 nm. The repetition rate of the laser was 1 kHz, yielding an optical fluence of 3.75 µJ/mm^2^, which is far below the safety limit according to the American National Standard for Safe Use of Lasers. An optically and acoustically transparent plastic membrane was affixed using surgical tape on the patient’s skin over the examined ROI. Both the laser output and ultrasound transducer (UT) were mount on the same scanning head placed close to the membrane to position the focal point of the ultrasound detector slightly above the skin surface and maximize detection sensitivity. The scanning head contained water as a coupling medium. Two mechanical stages (PI, Germany) were used to move the RSOM head. Both the laser and the controller of the mechanical stages were placed inside a plastic case, which ensured laser safety for all participants, as shown in (supplementary Fig. 1). The scanning field of view is 4×2 mm with a step size 7.5 µm in the fast axis and 15 µm in the slow axis.

### Recruitment, data grouping and statistical analysis

Eighty (N1 = 80) patients were scanned following approval from the local Ethics Committee of the Hospital of the Technical University of Munich. Besides diabetic patients, 20 healthy volunteers were scanned with matched ages of the patient group. All participants gave written informed consent. The detailed information of diabetic patients and healthy volunteers is shown in supplementary Table I. RSOM data quality was evaluated based on our previously developed RSOM quality evaluation approach and low-quality data was excluded. Finally, RSOM data from 72 patients and 20 volunteers were left for image reconstruction and further analysis. The patients were split into three main groups based on the complications of neuropathy and atherosclerosis. Group A consisted of 22 patients with no complications (neither neuropathy nor atherosclerosis). Group B included 25 patients with diabetic neuropathy but without atherosclerosis. Patients with diabetic peripheral neuropathy are evaluated by the Neuropathy Disability Score (NDS) and the Neuropathy Symptom Score (NSS). These scores are based on questionnaires and physical examinations, and measurements of sensory responses to temperature or vibratory stimuli ^35,36^. In order to quantify the effects of neuropathy, Group B was further divided into two sub-groups with the low NDS and NSS scores (n=13, 1 ≤ NDS ≤ 5 or 1 ≤NSS ≤ 5) and high scores (n=12, NDS > 5 or NSS > 5). Group C consisted of 25 patients with diabetic neuropathy and atherosclerosis.

**Table I:**
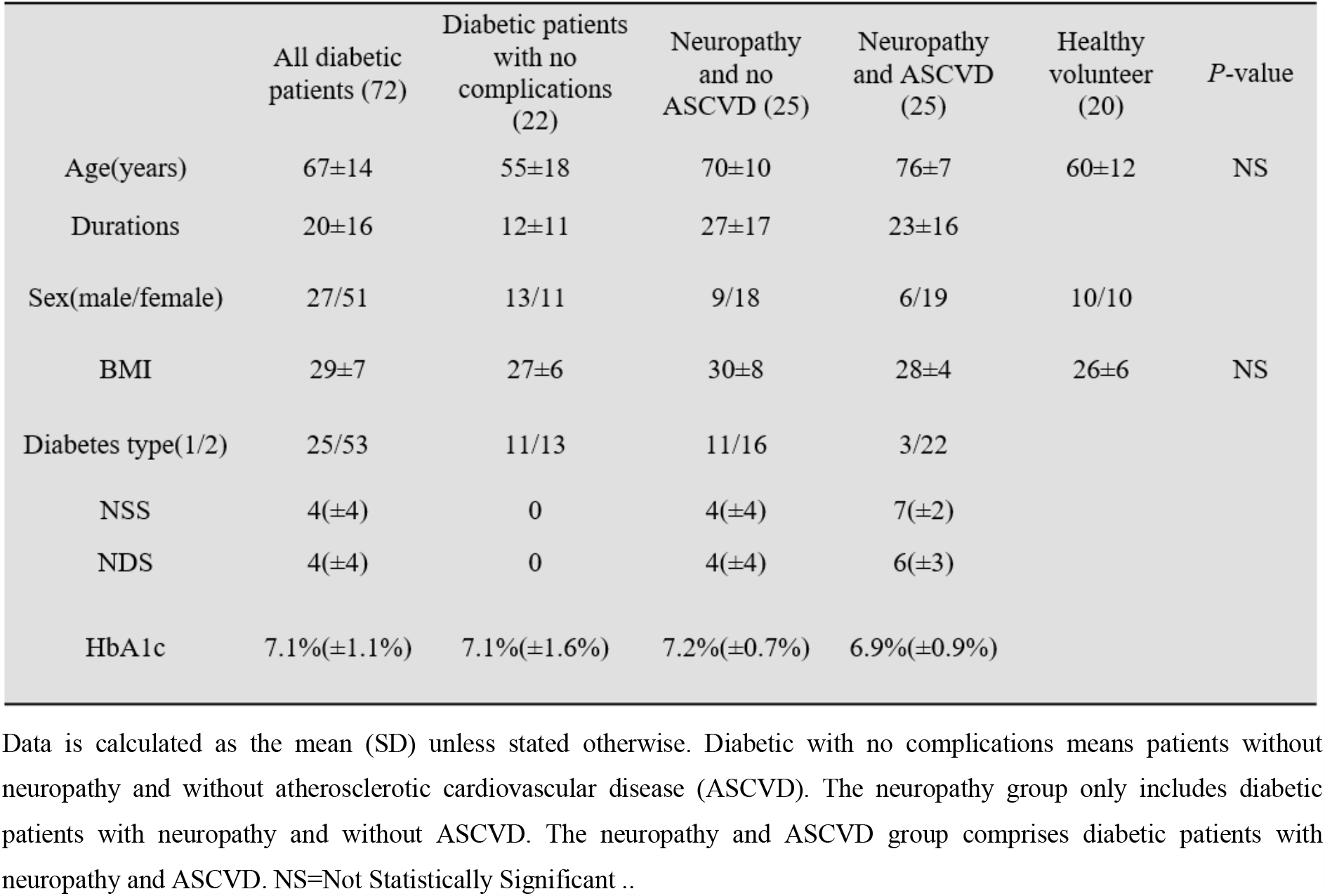
Characteristics of study participants

|  | All diabetic patients (72) | Diabetic patients with no complications (22) | Neuropathy and no ASCVD (25) | Neuropathy and ASCVD (25) | Healthy volunteer (20) | P-value |
| --- | --- | --- | --- | --- | --- | --- |
| Age(years) | 67±14 | 55±18 | 70±10 | 76±7 | 60±12 | NS |
| Durations | 20±16 | 12±11 | 27±17 | 23±16 |  |  |
| Sex(male/female) | 27/51 | 13/11 | 9/18 | 6/19 | 10/10 |  |
| BMI | 29±7 | 27±6 | 30±8 | 28±4 | 26±6 | NS |
| Diabetes type(1/2) | 25/53 | 11/13 | 11/16 | 3/22 |  |  |
| NSS | 4(±4) | 0 | 4(±4) | 7(±2) |  |  |
| NDS | 4(±4) | 0 | 4(±4) | 6(±3) |  |  |
| HbA1c | 7.1%(±1.1%) | 7.1%(±1.6%) | 7.2%(±0.7%) | 6.9%(±0.9%) |  |  |

Data is calculated as the mean (SD) unless stated otherwise. Diabetic with no complications means patients without neuropathy and without atherosclerotic cardiovascular disease (ASCVD). The neuropathy group only includes diabetic patients with neuropathy and without ASCVD. The neuropathy and ASCVD group comprises diabetic patients with neuropathy and ASCVD. NS=Not Statistically Significant ..

All metrics were displayed into column table with mean value and standard deviations as error bar. To assess the significance of the statistical differences for the metrics between healthy and diabetic patient groups, and sub-groups among diabetic patients, we performed parametric tests (unpaired t test) for normally distributed data; otherwise, nonparametric tests (Mann Whitney U test) were applied. Statistical significance was assumed at P < 0.05.

### Patient preparation and image acquisition

Patients were asked to consume no caffeine or food for at least 4 hours before the RSOM scans. They were placed in a quiet and dark room and left to relax for at least 5 minutes. Temperature of the room was held stable at 23°C during the whole procedure. Scans were performed with the patients in the supine position. Each patient was scanned at 2 symmetric regions of interest (ROIs, 4 × 2 mm) over the pretibial region of the distal lower limb. The scan of the dominant leg was used for further analysis. The pretibial region was used as representative of skin microcirculation, since the patients with diabetes are prone to developing cutaneous alterations at this very position. Each RSOM scan lasted approximately 70 sec. Before each scan the skin was cleaned with alcohol wipes. Both the patients and the operators used appropriate goggles for laser safety reasons.

### Image reconstruction

For the image reconstruction, acquired RSOM signals were divided into two frequency bands, 10-40 MHz (low) and 40-120 MHz (high), for the 10-120 MHz bandwidth. Signals in the two different bands were independently reconstructed. Reconstructions were based on beam-forming algorithms that generated three-dimensional images ^48^. The reconstruction algorithm was accelerated by parallel computing on a graphics processing unit, and improved by incorporating the spatial sensitivity field of the detector as a weighting matrix. The reconstruction time of one bandwidth takes about 5 minutes with voxel size of the reconstruction grid at 12 µm × 12 µm × 3 µm. The two reconstructed images *R*_*low*_and *R*_*high*_ corresponded to the low- and high frequency bands. A composite image was constructed by fusing *R*_*low*_into the red channel and *R*_*high*_ into the green channel of an RGB image. The detail process has been introduced in our previous work ^27^. The RSOM images can be rendered by taking the maximum intensity projections of the reconstructed images along the slow axis or the depth direction as shown in (Fig. 2).

### Quality Control

For this study, we recruited 80 patients and 20 healthy volunteers and recorded 200 RSOM measurements (two scans per person). Our previous studies have shown that motion can significantly affect image quality, although our motion correction algorithms can offer marked improvements ^49,50^. However, various motions from physiological displacements due to arterial pulsation and heartbeat and unintentional movements of the patient may lead to inconsistent motion correction improvements.

Therefore, we developed a quality control scheme based on the amount of motion in the raw data that classifies the quality of data collected. The quality control scheme enables the selection of high quality datasets, in which the motion is minimal enough for the motion-correction algorithm to correct, resulting in consistent correction improvements and uniform image quality for quantitative analysis ^50^. After the data quality evaluation, RSOM datasets of 8 patients were excluded due to serious motion and low image quality.

### Skin layer and microvasculature segmentation and calculation of RSOM-based biomarkers

For layer segmentation, RSOM images were first flattened based on our surface detection approach ^50^. The reconstructed volume of selected frequency band (10-40 MHz) was split into four stacks with 0.5 mm thickness along the slow scanning axis. Then, the epidermis layer in the MIP image of each stack was automatically segmented by a graph theory and dynamic programming based approach ^33^. The segmented boundaries of the epidermis layer from the four stacks were smoothed to achieve the final segmented results, as shown in (Fig. 1b and 1c). The thickness of the epidermis layer was calculated as the average width of the four segmented boundaries. Additionally, the signal density of the epidermis layer was determined as the ratio between the sum of the pixel intensity in the epidermis layer and the total segmented volume of the epidermis layer in the 4×2 mm scanning region.

The dermis layer was segmented starting from the bottom boundary of the epidermis layer and extending 1.5 mm deep. In the 4×2 mm^2^ scanning region, the total blood volume in the segmented dermis layer was calculated as ratio 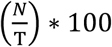, where *N* represents the number of voxels with intensities above 20% of the maximum voxel intensity, and *T* is the total voxel number inside the 4×2×1.5 mm^3^ volume. Afterwards, the vascular mask in the dermis was segmented by the multi-scale matched filter based vessel segmentation algorithm, as shown in (Fig. 1d) ^34^. Based on the mask, vessel boundaries were extracted and the corresponding width of the boundaries was calculated as the vessel diameter (Fig. 1e). Then, the centrelines of vessel boundaries were extracted and junction points of the centrelines were counted as the total vessel number (**see Suppl. Fig. S2**). To reduce noise or artefacts, we removed independent vessels with lengths less than 5 pixels (20 µm spatial resolution divided by 3 µm pixel size is approximately 7). The small vessel number was determined based on the number of junction points, where the average diameter of the connected vessel was less than 40 µm.

## Data Availability

Data is available from the authors upon receipt of a reasonable request. All patient data is confidential.

## Abbreviations

RSOM: Raster-Scan Optoacoustic Mesoscopy
OCT: Optical Coherence Tomography
MIP: Maximum Intensity Projections
ER: Epidermal
DR: Dermal
NSV: Number of Small Vessels
NLV: Number of Large Vessels
TVN: Total Vessel Number
TBV: Total Blood Volume
ET: Epidermal Thickness
ESD: Epidermal Signal Density
NDS: Neuropathy Disability Score
NSS: Neuropathy Symptom Score
NC: No Complications
LN: Low score Neuropathy
HN: High score Neuropathy
N_NA: Neuropathy and No Atherosclerosis
N_A: Neuropathy and Atherosclerosis

## Acknowledgements

This project has received funding from the European Union’s Horizon 2020 research and innovation programme under grant agreement No 687866 (INNODERM), from the European Research Council (ERC) under the European Union’s Horizon 2020 research and innovation programme under grant agreement No 694968 (PREMSOT) and from Helmholtz Zentrum München through Physician Scientists for Groundbreaking Projects, in part by the Helmholtz Association of German Research Center, through the Initiative and Networking Fund, i3 (ExNet-0022-Phase2-3). We thank Dr. Robert J. Wilson for his attentive reading and improvements of the manuscript and the staff at the Diabetes Center at Marienplatz in Munich and the Clinic of Vascular and Endovascular Surgery, Klinikum Rechts der Isar at TUM for assisting with the patient studies.

